# Impact of Shifting University Policies During the COVID-19 Pandemic on Self-Reported Employee Social Networks

**DOI:** 10.1101/2024.02.08.24302489

**Authors:** Stephanie S. Johnson, Katelin C. Jackson, Eric T. Lofgren

## Abstract

**Objectives:** To ascertain if faculty and staff were the link between the two COVID-19 outbreaks in a rural university county, and if the local university’s COVID-19 policies affected contact rates of their employees across all its campuses.

**Methods:** We conducted two anonymous, voluntary online surveys for faculty and staff of a PAC-12 university on their contact patterns both within and outside the university during the COVID-19 pandemic. One was asked when classes were virtual, and another when classes were in-person but masking. Participants were asked about the individuals they encountered, the type and location of the interactions, what COVID-19 precautions were taken – if any, as well as general questions about their location and COVID-19.

**Results:** We received 271 responses from the first survey and 124 responses from the second. The first survey had a median of 3 contacts/respondent, with the second having 7 contacts/respondent (p<0.001). During the first survey, most contacts were family contacts (Spouse, Children), with the second survey period having Strangers and Students having the most contact (p<0.001). Over 50% of the first survey contacts happened at their home, while the second survey had 40% at work and 35% at home. Both respondents and contacts masked 42% and 46% of the time for the two surveys respectively (p<0.01).

**Conclusion:** For future pandemics, it would be wise to take employees into account when trying to plan for the safety of university students, employees, and surrounding communities. The main places to be aware of and potentially push infectious disease precautions would be on campus, especially confined spaces like offices or small classrooms, and the home, as these tend to be the largest areas of non-masked close contact.

## Background

The COVID-19 pandemic has taken over a million American lives, and has left many more disabled since its arrival on American soil in early 2020(1). When the pandemic started spreading across the country, many higher education institutions shifted to online learning, and had most non-essential employees work from home (2–4). Universities swiftly postponed, cancelled, or severely limited all campus-related activities, including research, administration, exams, sports, and special events. If an activity could be done remotely (e.g., administrative affairs, office hours, committee meetings) this was by and large the only, or at the very least strongly preferred, route.

After the federal government declared the COVID pandemic a national emergency on March 13, 2020, public health experts and authorities recommended or imposed social isolation as a primary measure to ‘flatten the curve’ and mitigate the spread of SARS-CoV-2.(5) People of all ages were asked to avoid physical social contact, gatherings outside their immediate household, and to self-isolate even further if traveling. They were also recommended or mandated to wear a mask and separate up of six feet in public places (e.g. check out lines, restaurant tables)(6–8).

As the COVID-19 pandemic matured, economic worry and political considerations eroded public health measures. State education institutions guidelines and requirements were usually set by state health guidelines in addition to economic hardships – especially in regard to sporting events, on-campus housing, and teaching style- and as the state decided to lift measures, so would universities (9–11). This has led to universities teaching and staffing styles to change from virtual, to hybrid, to in-person – either with or without an accompanying mask mandate.

While the majority of attention on university campuses during these tumultuous school years has been on students, faculty and staff have also been affected by the shifting university policies. Faculty and staff tend to be the bridge between university students and the surrounding communities, especially in rural universities. While students tend to socialize with each other, employees enter and exit the community and university campus daily, and tend to have more community involvement (e.g local schools, community groups, etc.) than the students(12, 13).

Public health officials in Whitman County and Washington State University were able to track and count COVID-19 cases during the Fall 2020 semester, when classes were virtual, but a majority returning students came back to Pullman, giving it one of the highest cases per population(14, 15). There were two metapopulations noted within Whitman County, the students had a high peak and then a slight lull in August, where the non-student population then had a small outbreak in November(15). We were interested in trying to ascertain if faculty and staff were the link between the two outbreaks in Whitman County, and if Washington State’s COVID-19 policies affected contact rates of their employees across all its campuses. To answer these questions, we modified the FluScape survey used for the H1N1 influenza outbreak in China and Hong Kong for COVID-19 and asked university employees about their contacts during virtual learning period, and then in-person masking (16–18).

## Methods

The study protocol was approved by the Washington State University’s Institutional Review Board. The survey was modified from the FluScape Survey and administered through REDCap an electronic data capture tool hosted at Washington State University(19, 20).

Faculty and staff from all campuses of Washington State University were invited to participate in a contact tracing survey during two different survey periods. The first survey period was from January to March 2021, when the university had virtual learning and most employees were allowed to work from home. The second survey period was from November 2021 to March 2022, when the university was teaching in-person with a mask mandate and employees were being asked to go back into the office. Recruitment involved sending out listserv emails from the Office of Research, Deans of Colleges, and Department Heads with a link to the survey along with follow-up emails reminder emails. The respondents were kept anonymous and no personal demographic information was collected.

Both surveys sent out had three sections. The first section determined eligibility, defined as anyone over the age of 18, who was not a student, and was currently working for the university. In the second section, participants were asked to fill out a contact questionnaire, which aimed to record their physical contacts as they remembered them for the previous day. Physical contacts were defined as having a face-to-face conversation, skin-to-skin contact, indoor in a room for 10+ minutes, or non-socially distanced indoors or outdoors, for 10+ minutes. The encounter patterns of this study were in qualitative agreement with other contact studies, as well as initial evidence regarding COVID-19 aerosolization(17, 21–23). Respondents were asked to give a general label of a contact (e.g. colleague, barista, spouse), age range of contact, where they met with the contact (general location they wrote down and one of six general location types), how long they interacted, and if they touched or wore masks.

The third section asked general questions regarding the campus they worked at, their job title, if their job was classified as essential, and if they had had COVID-19 testing or symptoms since the beginning of the pandemic. Data was cleaned for general labels for contacts and specific locations. Cleaning and analysis was done with RStudio 4.2.2(24). The relationship between total contacts (degree) and potential variables that affected total contacts was evaluated using negative binomial regression. We express the output of these regression models as a degree ratio (DR), the ratio of counts compared to the referent group. Survey questions from both survey periods can be found in Supplemental Appendix A.

## Results

### Study Demographics Information

For the first period study, 413 respondents started the REDCap survey. 406 met eligibility requirements, and 271 respondents completed the contact questionnaire part of the survey. For the second study period, 323 respondents started the survey, 310 met eligibility requirements and 124 respondents completed the contact questionnaire (Figure 1).

**Figure 1.**
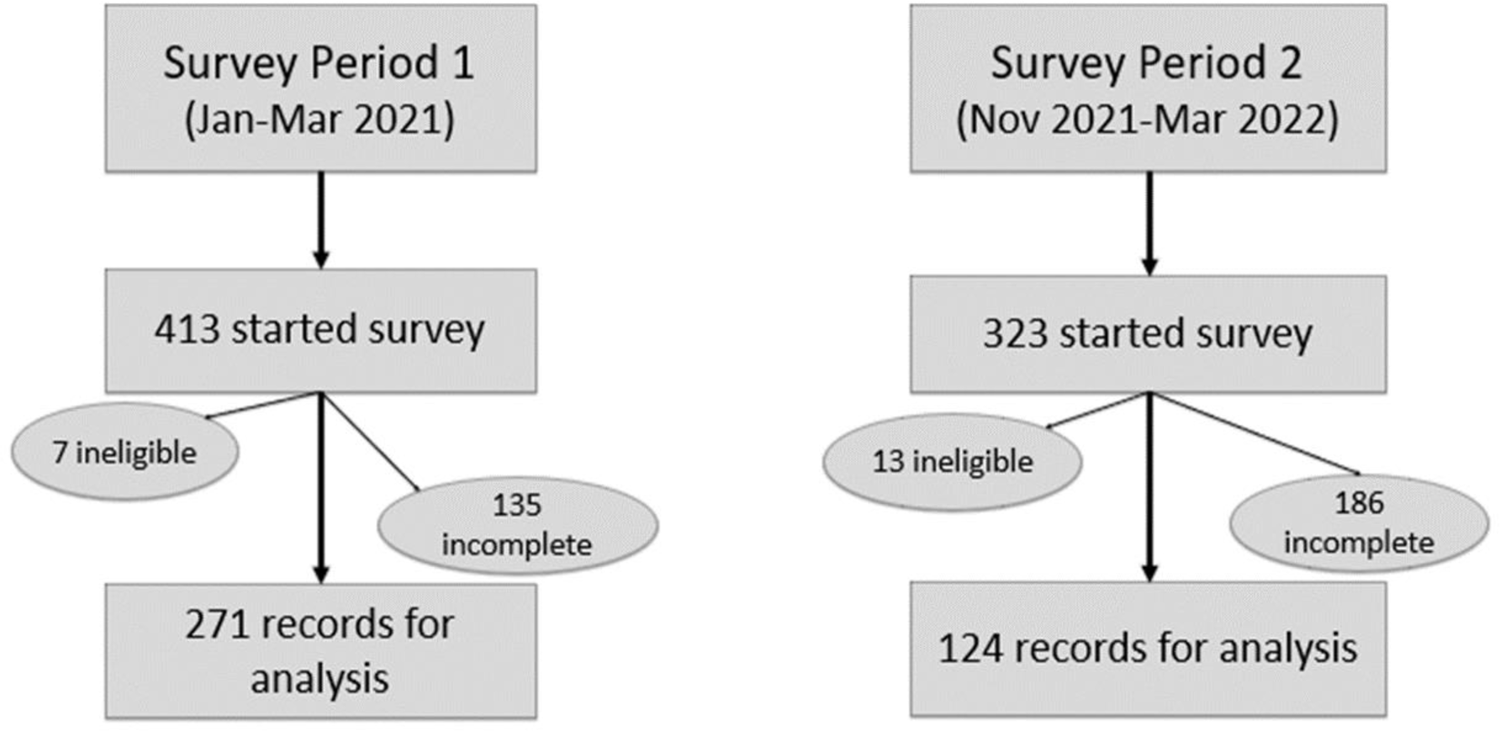
Flowchart for the study enrollment process for non-student university employees over eighteen years of age, and the resulting sample size for the virtual learning (Study Period 1) and in-person, masked instruction (Study Period 2) periods.

**Figure 2.**
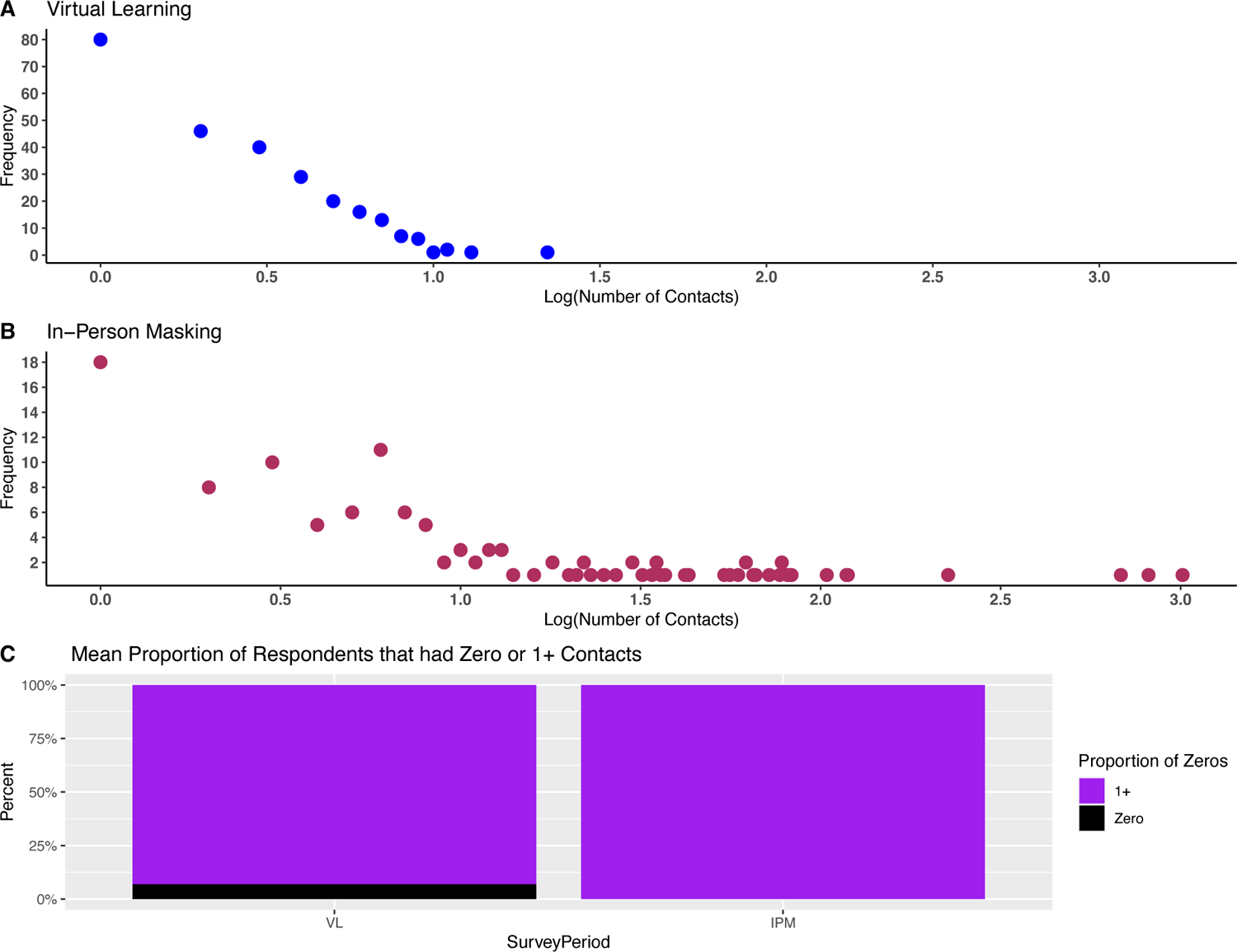
Degree distributions and proportion of surveyed respondents reporting zero contacts among university employees during virtual learning and in-person masked study periods respectively.

We break down the study period into Virtual Learning (VL) for January to March 2021, and In-Person Masking (IPM), for November 2021 to March 2022. We had 271 VL respondents with 872 contacts and 124 IPM respondents with 5024 contacts. Density histogram for the two study periods are shown in Figure2. The median contacts per respondent were 3 for VL and 7 for IPM (p <0.001)(Table 1). Most respondents during the VL period lived or worked on the main campus in Pullman WA (61%), while only 49% of respondents worked on that campus for IPM (49%)( χ^2^ =27.12, p <0.001). Further breakdown of occupation and essential workers between the two different learning periods are shown in Table 1.

**Table 1.** Median (IQR) Number of Recorded Contacts per Respondent per Day by Different Characteristics and Two Teaching Styles, Virtual Learning (VL), and In-Person Masking (IPM)

| Category | Covariate | VL Participants | Median (IQR) of Reported Contacts | IPM Participants | Median (IQR) of Reported Contacts |
| --- | --- | --- | --- | --- | --- |
| Total | --- | 271 | 3(3) | 124 | 7 (26.5) |
| Pullman (Live/Work) | Yes | 166 | 3(4) | 61 | 9 (27) |
|  | No | 56 | 2(3.25) | 55 | 3 (29.5) |
|  | NA | 49 | 2(2) | 8 | 2 (3.5) |
| Occupation | Faculty | 162 | 3(4) | 84 | 8 (32) |
|  | Staff | 96 | 2(3) | 27 | 7 (16) |
|  | NA | 13 | 2(3) | 13 | 5 (6) |
| Essential Worker | Yes | 61 | 4(5) | 56 | 12 (31.2) |
|  | No | 200 | 2(3) | 57 | 6 (13) |
|  | NA | 10 | 1.5(1.75) | 11 | 4 (5.5) |

Respondents were asked if they had ever been tested for COVID, and how many had been positive. 48 VL respondents had been tested in the last 30 days with no positives and IPM had 28 respondents tested in the last 30 days with 1 positive (χ^2^ =1.00, p=0.32). For total positives, VL had 10 respondents answer that they ever tested positive and IPM had 13 answer that they had ever tested positive for COVID(χ^2^ =5.98, p=0.015).

### Types of Contacts and Locations

When contacts were assessed, there was a different makeup for VL compared to IPM contacts. Most contacts in VL were family contacts, such as Partner (209) and Children (166), followed by Laborers (142). IPM had Strangers as the largest number of contacts (2639), followed by Students (1420), and then Colleagues (339) (χ^2^ =2183.3 p <0.001). (Figure 3a)

**Figure 3.**
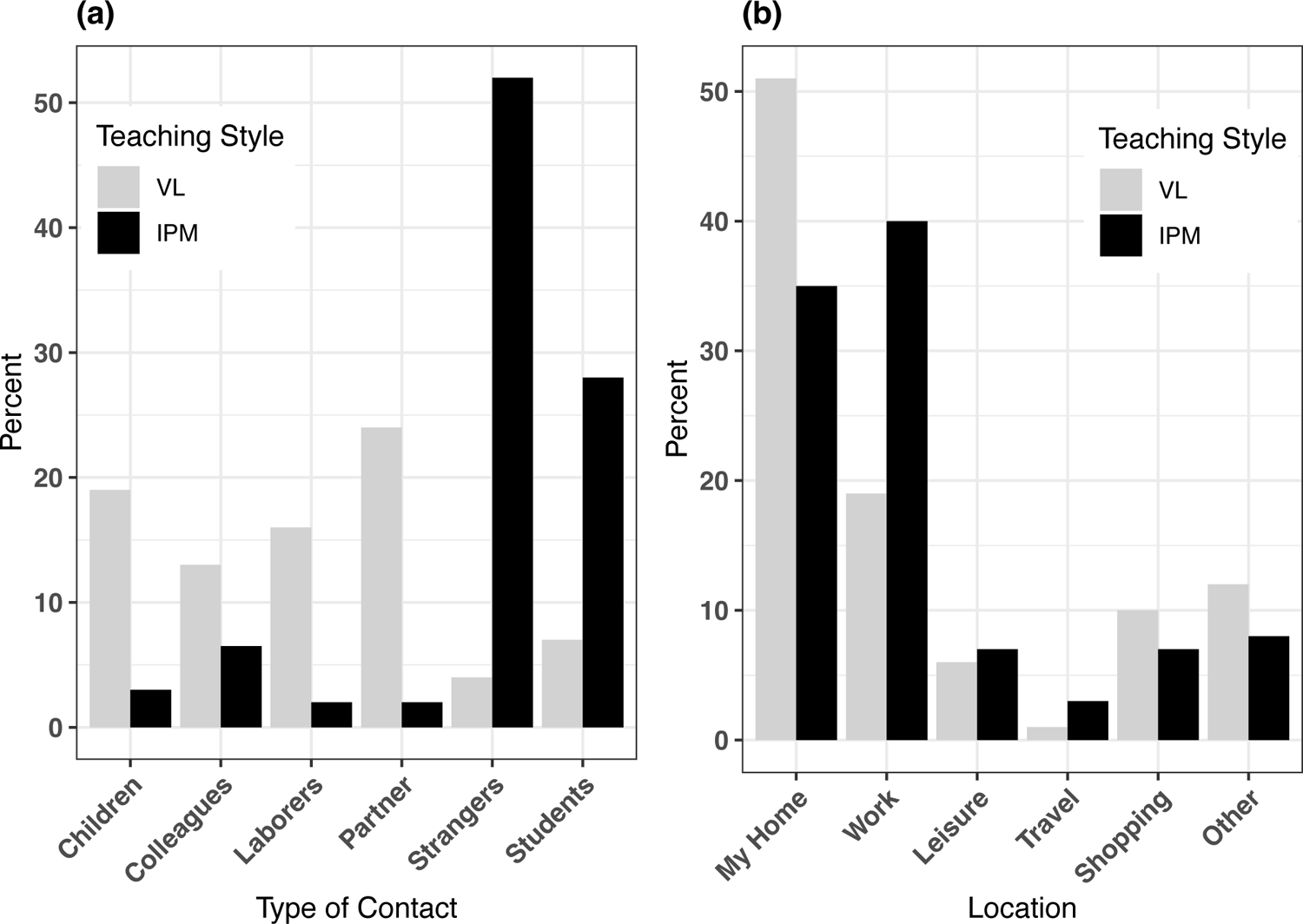
Distribution of type of contact by type of individual and location among university employees during virtual learning and in-person masked study periods respectively.

Respondents were able to choose one of six different places when asked where they had contact with people: My Home, Work, Leisure, Travel, Shopping and Other. This noted where contact happened, not how many people a respondent had contact with at each place. For VL, My Home was over 50% of responses, followed by Work (18%). IPM was a slightly more even split, with 40% of contacts happening at Work, and 35% at My Home. (χ^2^ =81.66, p <0.001) (Figure 3b)

### Masking

Respondents were asked about masking, as part of an awareness of how often they were masking. There were 44(5%) discordant pairs in VL and 47(10%) in IPM, where the respondent did the opposite masking as the contact. Neither party masked in 467 (53%) of VL encounters, and 227 (45%) of IPM encounters. Both parties masked 42% and 46% of the time for VL and IPM respectively (χ^2^ =13.6, p <0.01). The majority of masking was done with contacts they met outside of the house.

### Close Contacts

Close contact was defined as someone the respondent marked as having physical contact with. The top three types of close contacts were family members, specifically Partners/Spouses, Children, then other Family Members (e.g. grandparents, cousin, aunt). When analyzing close contacts by duration of interaction, it was noted that as duration of contact increased, so did the chance of the contact being a close contact. Contacts in VL were the ones most likely to be close contacts at shorter periods of time (χ^2^ =380.37, p<0.001). 30% of 30-60 minute interactions involved close contacts, compared to less than 20% for IPM. Approximately 20% of VL contacts were defined as close contacts, and about 13% were defined as such for IPM (χ^2^ =19.7, p<0.001). 75% of all My Home interactions were determined to be close contacts for both periods followed by Other (20%), and Leisure (10%).

The majority of close contact encounters for both periods were unmasked (94% and 92%). When looking at non-close contacts, 73% of VL encounters were masked (χ^2^ =470.82, p <0.001) and 63% of IPM encounters were masked ( χ^2^ =182.13, p <0.001)

### Negative Binomial Regression

Negative Binomial was used in multiple univariate models to determine if there were any variables that would affect the number of contacts a respondent would have for both survey periods. During the VL period having received flu shot significantly associated with having more contacts (DR 1.37 (95% Confidence Interval: 1.10-1.72)), and being an essential worker (DR 0.74 (95% CI: 0.6-0.91)) or having a typical day (DR 0.78 (95% CI: 0.61-0.99)) was associated with less contacts. For IPM, being an essential worker (DR 3.22 (95% CI: 1.88-5.54)), having a COVID-19 vaccination (DR 5.12 (95% CI: 1.73-12.03)), and teaching (DR 5.53(95% CI: 3.19-9.39)) was correlated with an increase in contacts. All results can be found in Table 2.

**Table 2.**
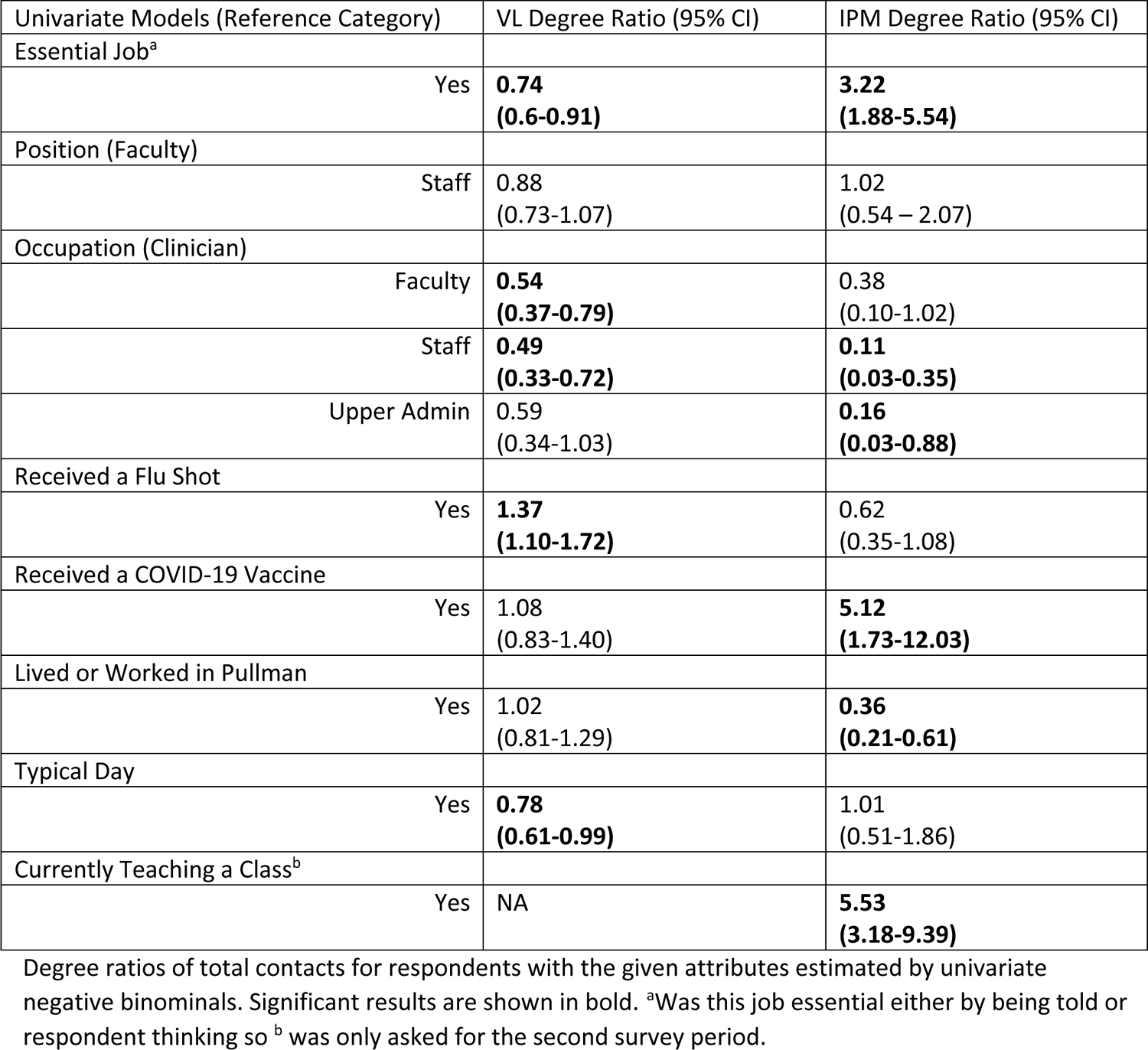
Univariate Negative Binomial Degree Ratios for both VL and IPM.

| Univariate Models (Reference Category) | VL Degree Ratio (95% CI) | IPM Degree Ratio (95% CI) |
| --- | --- | --- |
| Essential Job<sup>a</sup> |  |  |
| Yes | **0.74**<br>**(0.6-0.91)** | **3.22**<br>**(1.88-5.54)** |
| Position (Faculty) |  |  |
| Staff | 0.88<br>(0.73-1.07) | 1.02<br>(0.54 – 2.07) |
| Occupation (Clinician) |  |  |
| Faculty | **0.54**<br>**(0.37-0.79)** | 0.38<br>(0.10-1.02) |
| Staff | **0.49**<br>**(0.33-0.72)** | **0.11**<br>**(0.03-0.35)** |
| Upper Admin | 0.59<br>(0.34-1.03) | **0.16**<br>**(0.03-0.88)** |
| Received a Flu Shot |  |  |
| Yes | **1.37**<br>**(1.10-1.72)** | 0.62<br>(0.35-1.08) |
| Received a COVID-19 Vaccine |  |  |
| Yes | 1.08<br>(0.83-1.40) | **5.12**<br>**(1.73-12.03)** |
| Lived or Worked in Pullman |  |  |
| Yes | 1.02<br>(0.81-1.29) | **0.36**<br>**(0.21-0.61)** |
| Typical Day |  |  |
| Yes | **0.78**<br>**(0.61-0.99)** | 1.01<br>(0.51-1.86) |
| Currently Teaching a Class<sup>b</sup> |  |  |
| Yes | NA | **5.53**<br>**(3.18-9.39)** |

Degree ratios of total contacts for respondents with the given attributes estimated by univariate negative binominals. Significant results are shown in bold. <sup>a</sup>Was this job essential either by being told or respondent thinking so <sup>b</sup> was only asked for the second survey period.

## Discussion

For the first survey period, most of the respondents were on the main campus, and had very few contacts during VL. This corresponds with the lockdown that Washington State implemented as most employees or staff were accommodated with social distancing and work from home was encouraged or required. Bench lab work was performed in shifts, and those in teaching roles were mostly virtual (some health science classes were still in-person). VL also had the highest number of respondents marking their location with people as Other. When looking at the location that they wrote in, most of those were outdoor places-eg. parks, backyards, out on walks-showing that people were taking social distancing potentially more seriously in 2021, before vaccines against SARS-CoV-2 were introduced.

If a survey respondent indicated that they held an essential job during the VL period, they had a median of twice the number of daily contacts compared to non-essential respondents, which was not what the negative binomial analysis showed. When examining the essential worker contact distribution closer, a long tail of contacts appeared, from some faculty that still had to teach classes. This pattern persisted during the second survey period, where essential respondents experienced a more significant increase in contacts compared to non-essential respondents. We attribute this finding to essential people having more reasons for potential exposure, such as teaching large classes or participating in on-campus events. Additionally, the loosening of social distancing and mask-wearing practices over time during the ongoing pandemic played a role in this trend. This explanation is consistent with the observation that receiving a COVID vaccine during the second survey period was significantly associated with an increase in contacts. People likely felt safer with vaccination and began venturing out into public spaces more frequently. One of the essential workers for IPM mentioned going to a concert for the first time in two years. Results show how loosening policies and people going out in public had an impact using IPM’s highest contact category-Strangers. While VL had more people put down Shopping than IPM, IPM recorded a higher number of contacts at each location recorded.

When examining the incidence of close contacts across the two instructional policies, one can observe comparable levels of close contacts between VL and IPM. This similarity may be attributed to the consistency of contacts in close contact areas, suggesting that neither the easing of social distancing measures nor the increased presence of contacts elevated a respondent’s likelihood of close contact interactions with others. One’s home is still the place one will likely have the most amounts of close contacts, no matter the policies set forth by an institution or state. What was intriguing was the difference between close contacts and time spent between VL and IPM. The increased number of close contacts at shorter duration for VL might potentially speak of need for connection, or fewer number of people meeting in general, meant more of them were close contacts. With the sharp increase of total number of people IPM respondents have, they are more likely to touch those they are already close to and avoid touching others even if they are masked due to cultural norms and potential social distance guidelines.

There are caveats to this study. While the original survey period had a higher response rate, it was still less than 25% of employees across all the campuses, and samples heavily from faculty, rather than staff, who has 500 fewer appointees than faculty(25). Due to institutional concerns regarding confidentiality this study was unable to collect demographic information, negating opportunity to analyze contacts by age, gender, or ethnicity. This was also sent out via email, with many employees experiencing burnout or not having time to do a long study, as evidenced by the reduction of people who completed eligibility but not the contact diary. The study is susceptible to recall bias, as asking individuals to remember the details of a day within a 24-hour period is inherently challenging. These numbers are considered a conservative baseline, with the actual number of contacts, both close and otherwise, expected to exceed those reported here.

## Conclusion

This study is one of the few looking at university employees exclusively during two different policy periods that affected teaching, working, and learning for all campuses. Respondents were self-selecting and were able to offer an idea of what habits changed due to these institutional policies. We also must place results in context of what was going on during the time, not just policies but public opinion and other mandated health measures.

Notably, the interaction of respondents with students for the IPM survey response did not escape our attention. These were months where students were living on campus in large numbers, and classes were being taught in-person, with some potential disease mitigation measures(26, 27). University employees serving as a vital bridge between students and outside communities is a potential idea, especially in rural areas. In these regions there is more of a closed population and less chance for students to interact with strangers in large populations via public transit or population density(13, 28, 29). University employees have a unique position on college campuses, going back and forth as a liaison for students and the community. While employees might be teaching, advising, caring for, or helping students, they leave the campus and go out into the community and their own homes. They don’t live with the students in a mostly self-contained university-based social group but participate in both university and town ones.

For future pandemics, it would be wise to take employees into account when trying to plan for the safety of university students, employees, and surrounding communities. Employees tend to have a wide variety of contacts in multiple locations. The main places to be aware of and potentially push infectious disease precautions would be on campus, especially confined spaces like offices or small classrooms, and the home, as these tend to be the largest areas of non-masked close contact. The estimates from this study may assist in the planning of future campus-based interventions, predicting the shifts in social contacts that may be expected from a move toward virtual learning.

## Data Availability

All data produced in the present study are available upon reasonable request to the authors

## Acknowledgements

Funding: This work was supported by the Advancing Science in America Fellowship (Leo Barrier) and NIH R35GM147013. The authors would also like to acknowledge all the respondents who participated in these surveys.

